# Altered affinity to ACE2 and reduced Fc functional antibodies to SARS-CoV-2 RBD variants

**DOI:** 10.1101/2022.07.07.22277364

**Authors:** Ebene R Haycroft, Samantha K Davis, Pradhipa Ramanathan, Ester Lopez, Ruth A Purcell, Li Lynn Tan, Phillip Pymm, Bruce D Wines, P Mark Hogarth, Adam K Wheatley, Jennifer A. Juno, Samuel Redmond, Nicholas A Gheradin, Dale I Godfrey, Wai-Hong Tham, Kevin John Selva, Stephen J Kent, Amy W Chung

## Abstract

The emergence of severe acute respiratory syndrome coronavirus 2 (SARS-CoV-2) variants remains a formidable challenge to worldwide public health. The receptor binding domain (RBD) of the SARS-CoV-2 spike protein is a hotspot for mutations, reflecting its critical role at the ACE2 interface during viral entry. We comprehensively investigated the impact of RBD mutations, including 6 variants of concern (VOC) or interest (Alpha, Beta, Gamma, Delta, Kappa and Omicron) and 33 common point mutations, on IgG recognition, FcγR-engagement, and ACE2-binding inhibition in plasma from BNT162b2-vaccine recipients (two-weeks following second dose) and mild-to-moderate COVID-19 convalescent subjects using our custom bead-based 39-plex array. We observed that IgG-recognition and FcγR-binding antibodies were most profoundly decreased against Beta and Omicron RBDs, as well as point mutations G446S, found in Omicron, and N501T, a key mutation found in animal adapted SARS-CoV-2 viruses. Measurement of RBD-ACE2 binding affinity via Biolayer Interferometry showed all VOC RBDs have enhanced affinity to human ACE2. Furthermore we demonstrate that human ACE2 polymorphisms, E35K (rs1348114695), K26R (rs4646116) and S19P (rs73635825), have altered binding kinetics to the RBD of VOCs potentially affecting virus-host interaction and thereby host susceptibility.

## Introduction

Severe acute respiratory syndrome coronavirus 2 (SARS-CoV-2) — the virus responsible for causing Coronavirus Disease 2019 (COVID-19) — remains an ongoing challenge to global public health, as genomic surveillance continues to identify new mutations within the SARS-CoV-2 genome. These mutations have given rise to new variants of concern (VOCs), namely Alpha (B.1.1.7), Beta (B.1.351), Gamma (P.1), Delta (B.1.617.2) and most recently Omicron (B.1.1.529), each demonstrating increased transmissibility, leading to new waves of infections across the world (1, 2). Mutations to the receptor binding domain (RBD) of the SARS-CoV-2 spike protein are found across all VOC lineages and are of great interest (1). Binding of the SARS-CoV-2 RBD to angiotensin converting enzyme 2 (ACE2) receptor expressed on host cells facilitates virus entry and subsequent infection (3). As such, RBD mutations such as N501Y (found on both Beta and Omicron) which enhance the virus’ binding affinity to ACE2 can boost SARS-CoV-2 infection and transmission (4).

Antibodies play crucial roles in the body’s immunological response against SARS-CoV-2 infection. Neutralizing antibodies target the RBD and block engagement to ACE2 receptor on host cells, thus protecting from symptomatic infection (5, 6). Strong neutralizing activity against the ancestral RBD has been observed following seroconversion during SARS-CoV-2 infection, and COVID vaccination, particularly after receiving mRNA vaccines (such as Pfizer-BioNTech BNT162b2) (5). However, recent research has shown reduced neutralization capacity to VOCs, including Beta, Delta, and especially Omicron following both natural infection and vaccination (7–10). RBD mutations can result in a loss of epitope recognition by antibodies, such as the RBD mutation E484K (found in Beta and Gamma), which consequently reduces neutralizing capacity by antibodies (11). The recently emerged Omicron BA.2 harbours 16 amino acid changes at the RBD and has shown substantial escape from existing neutralizing antibodies (12). However, while RBD escape mutations can allow SARS-CoV-2 to evade neutralization, many of the antibodies elicited to the RBD can also induce non-neutralizing activity (13, 14).

Antibodies can coordinate additional functions via the crystallizable fragment (Fc) region, and growing evidence supports roles for Fc-mediated immunity in control of SARS-CoV-2 infection (15–18). Engagement of IgG with the IgG Fc gamma receptor (FcγR) on the surface of innate immune cells induces downstream effector functions, including antibody-dependent cellular phagocytosis (ADCP) (via FcγRIIa), and antibody-dependent cellular cytotoxicity (ADCC) (via FcγRIIIa) (19). ADCP has been described to occur following infection and vaccination in humans to ancestral SARS-CoV-2 (14). The importance of Fc-mediated immunity has been highlighted in animal studies, where animals that received neutralising monoclonal antibodies (mAbs) with compromised Fc-regions demonstrated greater viral burdens (15,18, 20). Despite the growing role for Fc-mediated immunity in SARS-CoV-2, the impact of RBD mutations on the capacity for antibodies to recruit these functions remains minimally described. Greater understanding of antibody Fc effector functions to VOCs would allow insights into potential protective signatures that remain following vaccination or infection across emerging viral variants.

Moreover, while mutations within the RBD can alter the virus’ affinity to ACE2, naturally occurring ACE-2 polymorphisms found within the human population, could also modulate host-pathogen dynamics during SARS-CoV-2 infection (21–24). Numerous human ACE2 (hACE2) protein-altering single nucleotide polymorphisms have previously been characterised with low frequency among the human population, including E35K (rs1348114695), K26R (rs4646116) and S19P (rs73635825) (22). ACE-2 polymorphisms which alter expression of ACE-2 on cells have also been predicted to affect affinity to RBD (21,22,24). The ACE2 polymorphisms S19P and K26R are predicted to confer increased susceptibility to infection due to increased binding affinity to ancestral SARS-CoV-2 RBD and full spike, as well as VOCS Alpha, Beta, Gamma (24–26). In contrast, the ACE2 polymorphism E35K has decreased binding to RBD constituting a more protective phenotype (24–26). However, the impact of RBD mutations of recently emerged VOCs, namely Delta and Omicron, on the affinity to these protein-altering ACE2 polymorphisms are yet to be investigated.

With the rise of VOCs, it remains crucial to understand the impact of RBD-mutations on key facets of host-interactions that may contribute to SARS-CoV-2 infection dynamics. Here, our study comprehensively measures the impact of 39-RBDs, including VOCs Beta, Omicron, and Delta, as well as common single point mutations, on the impact of IgG binding and ACE2 inhibition from the plasma of BNT162b2-vaccine recipients and mild-to-moderate convalescent donors using multiplexing. Our results show a loss of IgG -recognition against the RBD of VOCs, notably Omicron and Beta, and numerous single point mutations, including G446S and N501T. Importantly, this reduction translates to a loss of FcγR-binding antibodies and was validated using ADCP-assays, which demonstrate the capacity to induce Fc-effector functions are compromised against the RBD mutations found in VOCs. We also provide evidence that affinity between ACE2 and RBD influences the potential for antibodies to block ACE2-binding, using a surrogate *in-vitro* neutralisation assay. By using biolayer interferometry, we demonstrate that mutations found in VOCs and human ACE2 polymorphisms (E35K, K26R, S19P), alter the affinity and binding kinetics between RBD and ACE2, which may ultimately influence an individual’s susceptibility to COVID-19 infection.

## Methods

### Human Samples and Ethics Statement

Convalescent COVID-19 plasma samples were collected from individuals with mild-to-moderate disease as previously described (*n* =15) (27). Briefly, these samples were collected early-2020 (March to May) during the first wave in Australia and were likely infected with the ancestral strain. Vaccine plasma samples were obtained from individuals prior to vaccination (baseline) and two-weeks following second dose with Pfizer-BioNTech (BNT162b2) (*n* =16) as previously described (28). Participants characteristics for convalescent COVID-19 subjects and vaccine recipients are detailed in Table 1. Whole blood was collected into sodium heparin anticoagulant coated vacutainers before plasma was collected and stored at -80℃. Study protocols were approved by the University of Melbourne Human Research Ethics Committee (#2056689).

**Table 1:** Cohort Demographics

|  | **Baseline**<br>**(n = 16)** | **BNT162b2**<br>**recipients**<br>**(n = 16)** | **COVID-19 Patients**<br>**(n = 15)** |
| --- | --- | --- | --- |
| Age, median (IQR),<br>years | 30.5 [5.5] | 30.5 [5.5] | 49 [23] |
| Gender |  |  |  |
| Female, n(%) | 10 (62.5%) | 10 (62.5%) | 9 (60%) |
| Male, n(%) | 6 (37.5%) | 6 (37.5%) | 6 (40%) |
| Days from symptom<br>onset, median (IQR) |  |  | 38 [10.5] |
| Severity |  |  |  |
| Mild, n(%) |  |  | 9 (60%) |
| Moderate, n(%) |  |  | 6 (40%) |
| BNT162b2-vaccine |  |  |  |
| Days since<br>second dose,<br>median (IQR) |  | 13 [0.75] |  |

\* Mild defined as upper respiratory tract symptoms only; Moderate defined as including lower respiratory track symptoms. All patients non-hospitalised

### Proteins

SARS-CoV-2 antigens of the ancestral wild-type virus (B.1) are as follows: spike S1 (Sino Biological; 40591-V08H), spike S2 (ACRO Biosystems; S2N-C52H5), nucleoprotein (ACRO Biosystems; NUN-C5227) and spike trimer (provided by Adam Wheatley) (29). Influenza H1Cal2009 (Sino Biological; 11085-V08H) and SIVgp120 (Sino Biological; 40415- V08H) were used as positive and negative controls respectively (See Supplementary Table 1).

#### RBD Recombinant Proteins

SARS-CoV-2 RBD recombinant proteins, listed in Supplementary Table 2, were synthesized as previously detailed (30). Briefly, the sequences (obtained from GISAID surveillance repository) of the ancestral strain and RBD mutants were subcloned into pcDNA3.4 vectors by GenScript Corporation and subsequently expressed in Expi293 HEK cells (27). All RBD proteins underwent a two-step purification process, initially through HisTrap Excel HP columns (Cytiva) followed by a Superdex 75 Increase 10/300 pg gel filtration column platform (GE Healthcare), before protein concentrations were determined (absorbance measurement wavelength at 280nm). To study Omicron responses, ancestral WT RBD and Omicron BA.2 RBD were purchased from Sino Biological.

#### hACE2

Synthesis of truncated human ACE2 (residues 19-613 with a C-terminal AviTag and 6xHis-tag) was conducted as previously described (30). Targeted biotinylation was then performed using BirA enzyme prior to purification by strong anion exchange (Cytiva)(30).

### Multiplex Platform

#### Coupling of protein antigens to beads

Custom Luminex multiplex arrays were established to study SARS-CoV-2 antibody responses in accordance with previously established protocols (31). A complete list of protein antigen quantities-to-bead ratio for the *SARS-CoV-2-specific multiplex assay* and *ACE2- inhibition assay* can be found in Supplementary Table 1 and Supplementary Table 2 respectively.

#### SARS-CoV-2-specific multiplex assay

To characterize antibody responses across plasma samples, we utilized a broad panel of SARS-CoV-2 antigen coupled beads (see Supplementary Figure 1) in a Luminex multiplex array as previously described (Selva et al., 2021). Briefly, 1000 beads/bead region diluted in PBS containing 0.1% BSA (incubation buffer) were added to each well (in a total of 20µl volume/well) in 384-well plates (Greiner Bio-One; 781906), followed by addition of 20µl/well of plasma diluted at a single concentration (1:100 in PBS). Plates were incubated on a plate-shaker overnight at 4℃ before wells were washed with PBS 0.05% Tween-20 (PBST). Phycoerythrin (PE)-conjugated mouse anti-human antibodies that detect isotype (*Pan*-IgG, IgA1; Southern Biotech; 9040-09; 9130-09), or, biotinylated IgM (MabTech; mAb MT22), or, biotinylated Fc-receptors (FcγRIIa-H131, FcγRIIIa-V158) (kindly provided by Bruce Wines and Mark Hogarth (32)) were diluted to 1.3µg/ml and added at 25µl volume/well before 2hrs at room temperature on a plate shaker. For biotinylated detectors, plates were followed with an additional PBST wash, and incubated with streptavidin-PE (Invitrogen; S866) at room temperature for 1hr. Plates were then washed with PBST and each well resuspended in 50µl of sheath fluid. The level of PE-signal associated with each bead region in each well, reported as median fluorescence intensity (MFI), was determined by a Flexmap3D Luminex platform. Each sample was run in duplicate.

#### ACE2-inhibition assay

We employed a recently established surrogate neutralisation assay capable of simultaneously measuring the neutralising capacity of antibodies against multiple RBD variants (See Supplementary Table 2), following a slightly modified version of the protocol (30). Briefly, 700 beads/bead region diluted in incubation buffer were added to each well (in a total of 20µl volume/well) in 384-well plates, followed by application of plasma, prepared as an 8-point serial dilution beginning at 1:5 in PBS for each sample (20µl/well). Plates were covered and incubated on plate shaker for 1hr, and then 20µl/well of biotinylated-hACE2 diluted to 25µg/ml was added, followed again by 1hr shaking at room temperature. Plates were then washed with PBST and incubated with 40µl of streptavidin-PE (Invitrogen; S866) diluted to 4ug/ml in incubation buffer on a plate shaker for 1hr. Following incubation, plates were incubated for an additional 1hr with 10µl/well of R-PE Biotin-XX conjugate (Thermo Fisher Scientific; P811) diluted to 10µg/ml before washing with PBST and resuspension in 50µl of sheath fluid. Flexmap3D Luminex platform was used for acquisition of samples and reported as MFI for each bead region used. Plates were run in duplicate.

For wells used in the detection of isotype (IgG, IgM, IgA) and FcR-binding features of RBD-specific antibodies, 20ul/well of incubation buffer-*only* was added in replacement of diluted-biotinylated hACE2. Plates were then washed with PBST and incubated with respective detectors in methods described above.

### Biolayer interferometry (BLI)

The WT ACE-2 – RBD variant binding kinetics Bio-layer interferometry (BLI) was performed on the Octet Red instrument (FortéBio) as previously described (30). Briefly, Streptavidin Biosensors (FortéBio) hydrated in HBS-EP buffer ((0.01 M HEPES, 0.15 M NaCl, 3 mM EDTA, and 0.005% v/v Surfactant P20, pH 7.4 (GE Healthcare)) for 20 minutes were programmed to perform loading of biotinylated ACE-2 at 3µg/ml. Once the ACE-2 binding response reached a threshold of 1.6nm, the sensors were equilibrated in buffer-*only* wells for 120 seconds (baseline signal). The ACE-2 loaded sensors then captured respective RBD variants at concentrations ranging from 100nM to 6.25nM (diluted via 2-fold serial dilution) for 150s (association phase). Sensors were finally immersed in buffer-*only* wells for 360s to record the dissociation signal. Curve fitting was performed using a global fit 1:1 langmuir binding model using Octet Data Analysis software v12.0.2.3 (FortéBio), and baseline drift was corrected by reference subtracting the shift of an ACE2-loaded sensor immersed in kinetic buffer only. Mean kinetic constant values from 2 independent experiments were determined, with all binding curves matching the theoretical fit with an r2 value of more than 0.99. The complex t1/2 in seconds was calculated using the formula: t1/2 = 1n2/kdis = approximately 0.69/ koff.

BLI for measuring ACE-2 polymorphism binding affinity to RBD variants utilized the same protocol as above. The difference is only at the association phase, where sensors loaded with respective ACE-2 variants at 3ug/ml concentration were immersed into wells with RBD variants at a single concentration of 100nM for 150s. Baseline, loading and dissociation phases were as described above. A local fitting 1:1 binding model was used to calculate the curve fit and kinetic constants.

Both assays were performed in 96-well plates (Greiner Bio-One™ Polypropylene 96- Well F-Bottom Microplates, black (Interpath 655209) at 1000rpm and 30°C.

### Bead-based ADCP duplex assay

To assay the capacity for vaccinee plasma to mediate ADCP to SARS-CoV-2 wild type and to VOC’s, a competitive, sample sparing duplex ADCP assay was adapted from previously described assays (14,33,34). SARS-CoV-2 RBD wild type (WT) and SARS-CoV-2 RBD derived from the VOC Beta (B.1.351) were biotinylated using EZ-Link Sulfo-NHS-LC biotinylation kit (Thermo Scientific) with 20mmol excess according to manufacturer’s instructions and buffer exchanged using 30kda Amicon centrifugal filters (EMD millipore) to remove free biotin. Red or yellow-green 1μm fluorescent NeutrAvidin Fluospheres beads (Invitrogen) were coupled overnight at 4°C to RBD WT and RBD Beta respectively (1ul beads:3ug antigen). Coupled beads were washed thrice with 2% BSA/PBS to remove excess antigen, diluted 1:100 and combined to form a bead cocktail. 10ul of bead cocktail (9x10^5^ beads) was added to each well in a 96-well U-bottom plate with titrations of baseline and vaccinee plasma (1:100-1:777600) and incubated for 2 hours at 37°C. THP-1 monocytes (100,000 cells/well) were added to opsonized beads and incubated for 16 hours under cell culture conditions. Cells were fixed with 2% formaldehyde and acquired by flow cytometry on a BD LSR Fortessa with a HTS. The data was analyzed using FlowJo 10.8.1 (See Supplementary Figure 1 for gating strategy) and phagocytosis scores were calculated for each bead type (RBD WT and RBD Beta) as previously described using the formula: [%bead-positive cells × mean fluorescent intensity] (35). To account for non-specific uptake of S-conjugated beads, the phagocytosis scores for each plasma sample were subtracted with that of the “no plasma” control. ED50’s was calculated for each sample and assays were performed in duplicate.

### Statistics

#### Systems Serology (LASSO PCA)

Antibody features were selected using a least absolute shrinkage and selection operator (LASSO) reduction method as previously described using MATLAB version 9.6 (including machine learning and statistical toolbox) (The MathWorks, Inc., Natick, MA) and Eigenvector PLS toolbox (Eigenvector, Manson, WA) (36). In brief, data was first processed by right-shifting (to remove negative values) and log transformation (using equation log10 (x +1)) prior to being normalized using *z*-scoring. LASSO feature reduction was then employed on *z*-scored data and performed 1,000-times with 10-fold cross validation. Using LASSO-selected features, unsupervised principal component analysis (PCA) was then performed.

#### Data Analysis

Prism GraphPad version 9.0.2 (GraphPad Software, San Diego, CA) were used to develop graphs and perform statistical analysis as described in respective figure legends.

## Results

### Greater IgG and FcγR-binding antibodies to ancestral SARS-CoV-2 spike and RBD in BNT162b2 recipients compared to mild-to-moderate convalescence

Sixteen individuals vaccinated with BNT162b2 had plasma samples drawn before vaccination (baseline) and two weeks (median: 13 days; IQR: 0.75 days) following the second dose (Table 1). We also studied plasma from fifteen convalescent individuals (median: 38 days post symptom onset; IQR: 10.5 days) with mild-to-moderate COVID-19 disease. Importantly, convalescent samples were collected during the first wave of the pandemic (between March 2020 and May 2020 in Victoria, Australia), providing an opportunity to study responses in individuals infected by pre-VOC ancestral viruses that harbor less phylogenic deviation from the ancestral reference strain used in the BNT162b2 mRNA vaccine.

First, we characterized the plasma humoral profile to SARS-CoV-2 proteins (NP, RBD, S1, Trimer Spike) found in the ancestral wild-type virus. Using a SARS-CoV-2 multiplex bead assay, we measured antibody isotype (IgG, IgA1, IgM), and subclass (IgG1, IgG2, IgG3, IgG4) levels, as well as the capacity for antigen-specific antibodies to engage soluble FcγRIIa and FcγRIIIa-dimer constructs, which mimic the surface of innate immune cells and are a proxy readout for ADCP and ADCC activity respectively (19). Robust antibody responses to NP were uniquely associated with convalescent COVID-19 individuals, indicating no prior infection in the BNT162b2-vaccinees (Figure 1A; Supplementary Figure 2). Convalescent and BNT162b2- vaccinees both elicited elevated levels of IgG to spike antigens, however BNT162b2-vaccinees demonstrated significantly greater responses in comparison to convalescent individuals which displayed more heterogenous responses (S1, 3.8-fold higher; trimer, 1,6-fold higher; RBD, 3.5- fold higher), (Figure 1B-D; Supplementary Figure 2). Consistent with this, plasma from BNT162b2-vaccinees showed significantly higher FcγRIIa and FcγRIIIa-dimer binding levels by anti-spike antibodies than mild-to-moderate convalescent patients (Figure 1B, D; Supplementary Figure 2). We then used systems serology to broadly examine this repertoire of antibody responses and observed with principal component analysis (PCA) that the baseline, BNT162b2 (2-weeks post dose two), and convalescent (mild-to-moderate) subjects naturally separated into three distinct groups based on their antibody signatures (Figure 1E). A total of fifteen antibody features, selected using a LASSO (least absolute shrinkage and selection operator) feature-reduction tool, were key contributors to the differentiation between groups (Figure 1F). This analysis highlights that BNT162b2-vaccination induces more robust anti-ancestral spike IgG responses with greater FcγR-binding capacity, in comparison to mild-to-moderate convalescent donors.

**Figure 1.**
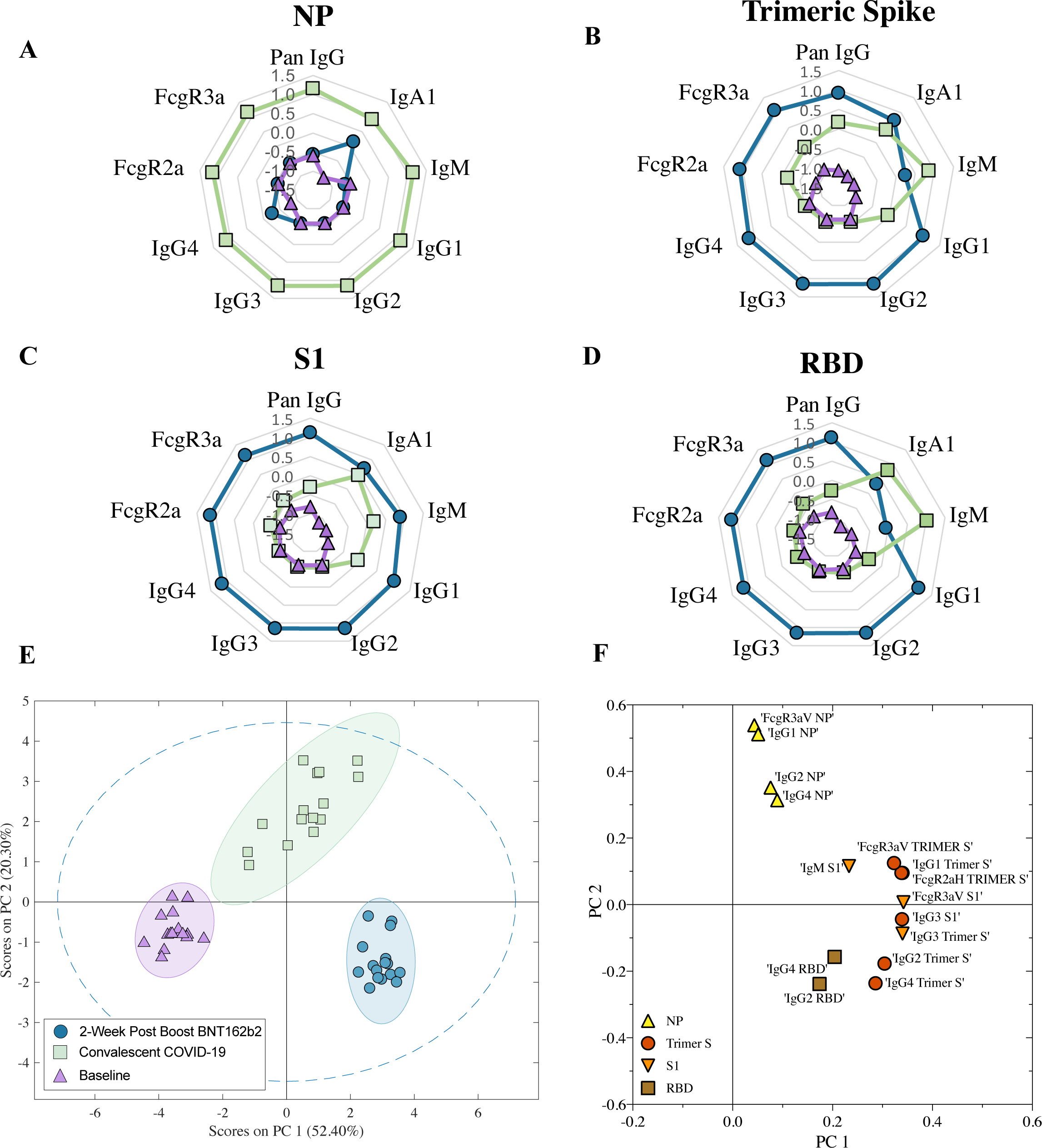
SARS-CoV-2 serological signatures distinguish BNT162b2 vaccinated, convalescent, and baseline individuals. Plasma humoral responses were profiled via multiplex for convalescent mild/moderate COVID-19 patients (green square; median 38 days post symptom onset; *n* = 15) and BNT162b2-vaccinated individuals at baseline (purple triangle; *n* = 16) and two-weeks following second dose (blue circle; *n* = 16). Radar plots show the median MFI (median fluorescence intensity) value of each antibody signature (IgG, IgA, IgM, IgG1-IgG4, FcγR- dimer binding) to NP (A), trimeric spike (B), S1 (C), and RBD (D). The median MFI for each feature was normalised using z-scoring prior to plotting. (E) Unsupervised principal component analysis comparing convalescent COVID-19 individuals and BNT162b2-vaccinated individuals at baseline and two-weeks following second dose of 15-SARS-CoV2 antibody features selected by a dimensionality reduction method (LASSO). 95% confidence intervals for each group are depicted by a coloured eclipse. (F) PC1 and PC2 loadings of LASSO selected features for NP (yellow triangle), trimeric spike (red circle), spike subunit-1 (S1) (orange inverted triangle) and receptor binding domain (RBD) (brown square) of the PCA.

### Reduced IgG binding antibodies to SARS-CoV-2 RBD variants, but not always concordant to loss in ACE2 inhibition

The RBD of the spike protein is a mutational hotspot for SARS-CoV-2. Because of its position at the ACE2 interface, mutations in the RBD can alter ACE2 affinity as well as result in the loss of epitopes for antibodies (2,37,38). Thus, to characterize the impact of RBD-mutations on plasma IgG binding in our convalescent and BNT162b2 donors, we generated a custom panel of 39 RBDs, including RBDs of 33-common point mutations, 5 variants of concern/interest (Beta, Delta, Gamma, Kappa, and Omicron) and the ancestral RBD, and measured antibody responses using multiplex. Higher levels of IgG binding responses (larger reciprocal ED50 values) were observed in BNT162b2-vacinees compared to convalescent individuals across all RBDs assessed (Figure 2A, E), consistent with our previous observation that BNT162b2 elicits more robust anti-spike IgG responses. Despite the overall greater levels of IgG in vaccinees, significantly reduced IgG recognition was observed against the VOCs Beta (7.1-fold reduction, p < 0.0001)) and Gamma (15.5-fold reduction, p < 0.0001) by BNT162b2 plasma in comparison to the ancestral RBD (Figure 2C; Supplementary Table 5).

**Figure 2.**
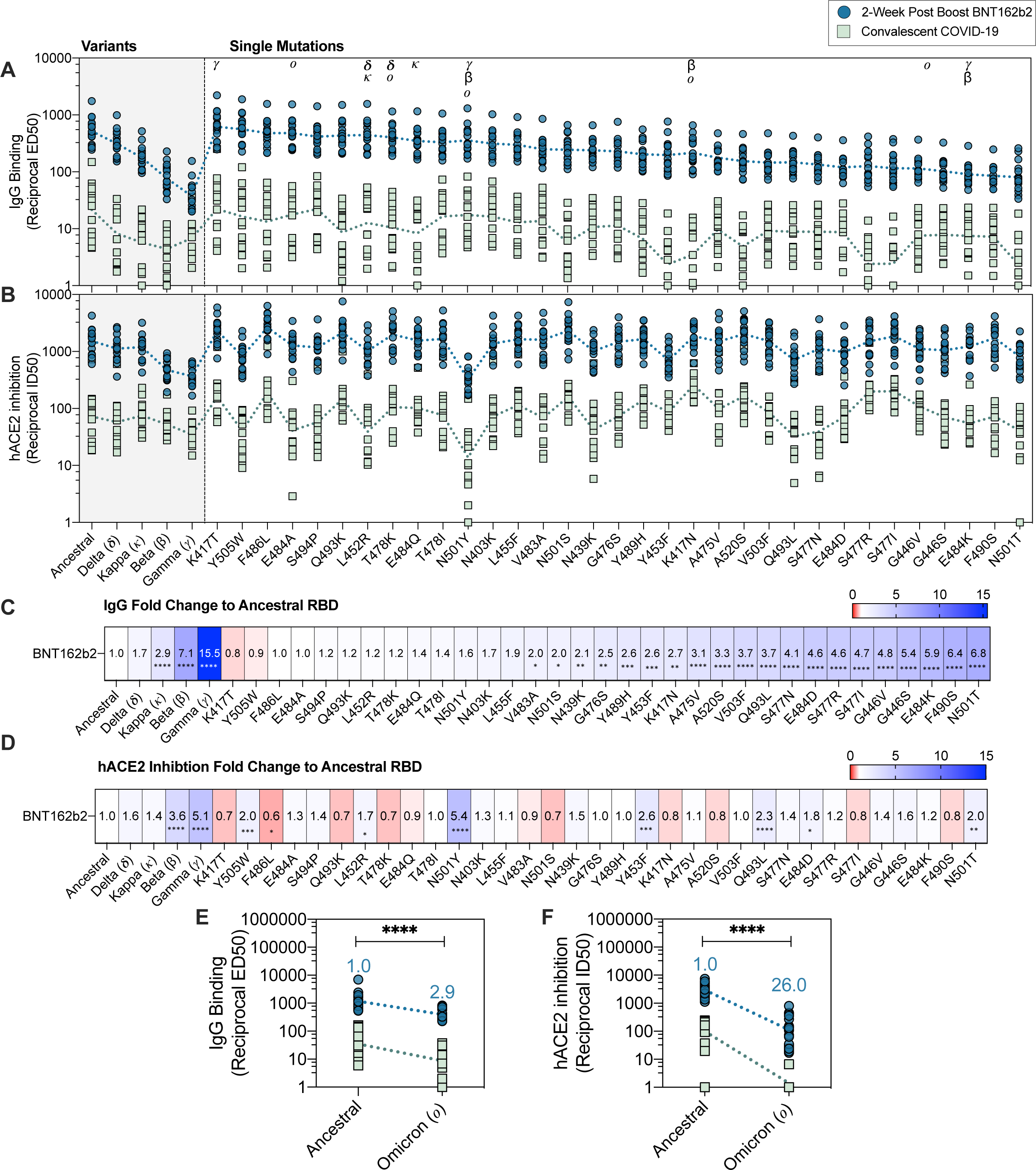
IgG-binding and hACE2 inhibition to ancestral RBD and 38-RBD variants. (A) *Pan*-IgG binding levels (reciprocal ED50) and (B) human wild-type ACE2 (hACE2) inhibition levels (reciprocal ID50) by BNT162b2 (two-weeks following second dose; blue; *n* = 16) and mild/moderate convalescent COVID-19 patients (green; median 38 days post symptom onset; *n* = 15) as measured by multiplex to RBD variants. Reciprocal ED50/ID50 values respectively were calculated using the normalized binding MFI-value for an 8-point serial plasma titration. (C) Fold change of *pan*-IgG binding levels of RBD variants in comparison to ancestral-RBD and (D) Fold change of hACE2 inhibition of RBD variants in comparison to ancestral-RBD by BNT162b2 plasma. Fold change was calculated as follows: (geometric mean of reciprocal ED50 or ID50 Ancestral-RBD) divided by (geometric mean reciprocal ED50 or ID50 variant-RBD). Friedman’s non-parametric test with Dunn’s multiple comparisons was used to assess statistical significance. (E) Pan-IgG binding levels (reciprocal ED50) and (F) hACE2 inhibition levels (reciprocal ID50) to RBD of Omicron BA.2 variant with fold-change as indicated above in blue. Mann-Whitney U-test used for statistical evaluation. *p* < 0.05 (*), *p* < 0.01 (**), *p* < 0.001 (***) and *p* < 0.0001 (****).

Drop in IgG binding was also observed with Omicron (BA.2) (2.9-fold reduction, p < 0.0001) (Figure 2E). Furthermore, significant decreases in IgG recognition to various point mutations, including G446S (5.4-fold reduction, p < 0.0001) found in Omicron, E484K (5.9-fold reduction, p < 0.0001) found in Gamma and Beta, as well as F490S (5.4-fold reduction, p < 0.0001) and N501T (6.8-fold reduction, p < 0.0001) (Figure 2C; Supplementary Table 5). This data highlights the influence of particular point mutations on recognition of the RBD by IgG antibodies.

Since the RBD is a dominant target for neutralizing antibodies, we next considered the impact of these RBD-variants on blocking ACE2 binding, using a surrogate neutralization assay. Using a previously established *in vitro* ACE2 inhibition assay, we measured the capacity for BNT162b2 and convalescent plasma to block hACE2 binding in a competitive format (30). Overall, the capacity for BNT162b2 plasma to block ACE2 binding was greater (larger reciprocal ID50 values) in comparison to convalescent plasma across all 39-RBDs (Figure 2B, F). Consistent with the loss of IgG binding, we observed decreased ACE2 inhibition to all assessed VOC, with a significant decrease observed for Beta (3.6-fold reduction) and Gamma (5.1-fold reduction), though this reduction was notably smaller than that for IgG binding (Figure 2C). A substantial drop in ACE2 inhibition was observed with Omicron (BA.2) (26- fold reduction; p < 0.0001) highlighting the evasive nature of this variant (Figure 2F). On the other hand, most of the RBD point mutations which displayed reduced IgG binding did not necessarily translate to a significant drop in ACE2 inhibition (Figure 2C, D). While a decrease in IgG binding was observed for the RBD mutations G446S and N501T, the loss in ACE2 inhibition observed was much weaker (G446S=1.6-fold reduction, p = ns; N501T=2.0-fold-reduction, p = 0.0013) (Figure 2D; Supplementary Table 3). A 1.7-fold-reduction (p = 0.04) in ACE2-blocking was also observed for L452R, despite having comparable IgG binding (p = ns) to the ancestral RBD. Furthermore, the RBD mutation N501Y, the prime mutation found in the RBD Alpha-VOC, demonstrated no significant decrease in IgG binding (1.6-fold reduction, p > 0.99) but showed a significant drop in capacity to block ACE2 from binding (5.4-fold reduction, p < 0.0001) (Figure 2C, D; Supplementary Table 5). Together, this data highlights the influence of naturally occurring RBD-mutations on the potential for polyclonal antibodies to recognize and block ACE2 binding.

### SARS-CoV-2 VOC RBDs have improved affinity to WT-hACE2

Mutations within the RBD also have the potential to modulate affinity to hACE2 (2,4,38,39). Hence, we next characterized the binding affinity of wild type (WT)-ACE2 to VOC RBDs, as well as a subset of single-point mutations, using biolayer interferometry (Figure 3; Supplementary Figure 3). We found all VOC RBDs assessed demonstrated elevated binding affinity to WT-ACE2 in comparison to ancestral-RBD (Kd: 29.4 nM; t1/2: 83.6 s) (Figure 3). These included Beta (Kd: 14.6 nM; t1/2: 164.83), Gamma (Kd: 15.0 nM; t1/2: 214.82 s) and Delta (Kd: 21.5 nM; t1/2: 143.77 s). Omicron (BA.2) demonstrated a 2.6-fold increase in binding (Kd: 11 nM; t1/2: 161.75 s), compared to ancestral-RBD (Figure 3). Together, these data reiterate how these mutations in the RBD, given its position at the ACE2-interface, can potentially improve viral fitness by enhancing affinity to ACE2. Furthermore, these data suggest that the RBD mutations observed in Beta and Gamma not only significantly reduced IgG binding, but also compounded with stronger ACE2 affinity to give the largest drops in ACE2 inhibition.

**Figure 3.**
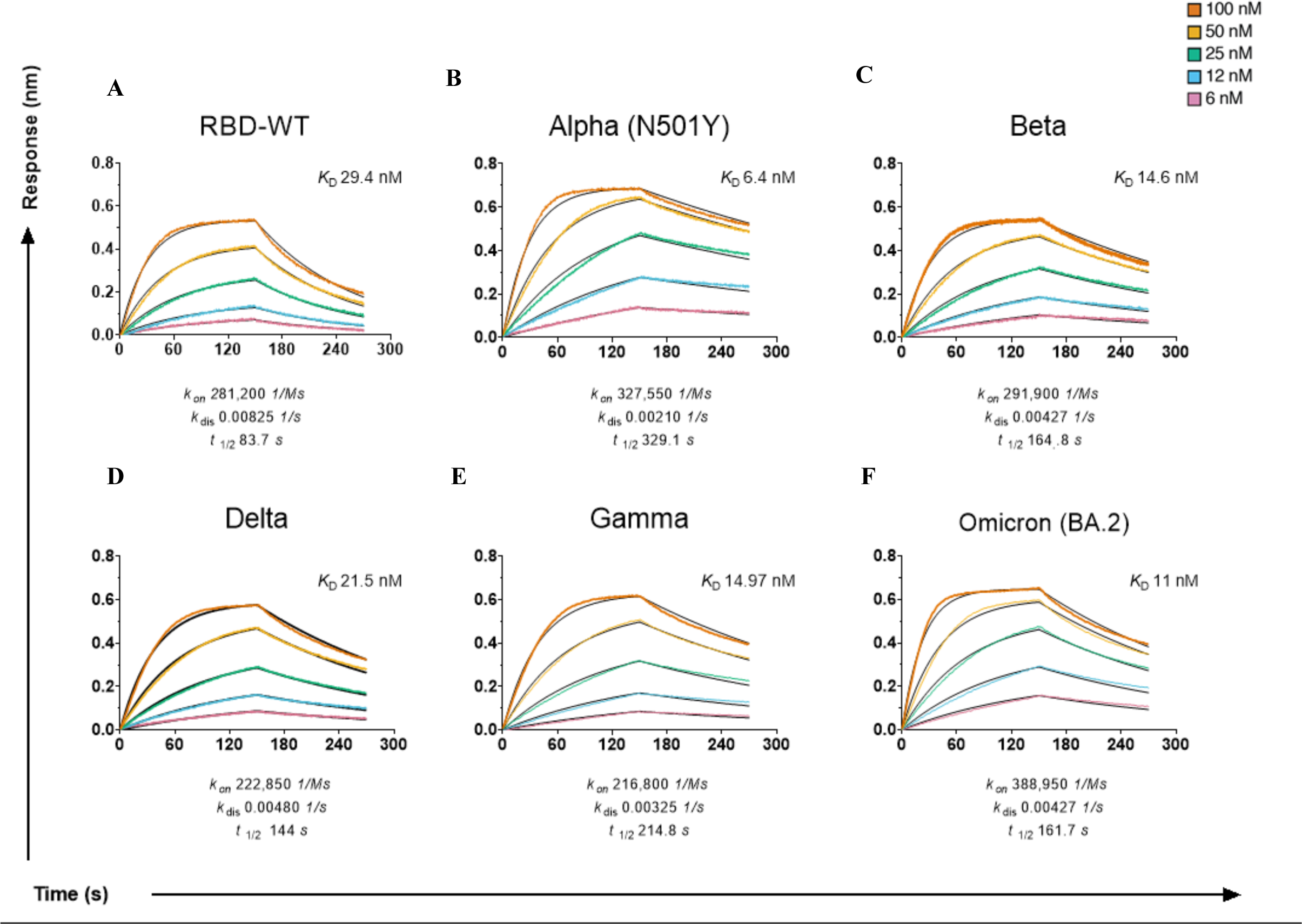
Affinity of wild-type (WT) hACE2 to the RBD of variants. Sensograms of WT-human ACE2 (hACE2) binding affinity to (A) ancestral-RBD, (B) Alpha (B.1.1.7), (C) Beta (B.1.351), (D) Delta (B.1.617.2) (E) Gamma (P1), (F) Omicron (BA.2) as measured by biolayer interferometry (BLI). Binding of 2-fold serial dilutions of each SARS- CoV-2 RBD variant (100 nM (red), 50 nM (yellow), 25 nM (green), 12 nM (blue), and 6 nM (pink)) was measured to immobilised hACE2 (3ug/ml). Binding curves representative of 2 independent experiments for each variant are plotted (solid-colored lines), globally fitted to a 1:1 binding model (black line).

Previously, we and others have shown single-point mutations in RBD (such as N501Y found in Alpha, Beta, Gamma and Omicron) can enhance affinity to ACE2 (4.5-fold increase; Kd: 6.5 nM; t1.2 : 329.1 s, Figure 3B) (4,30,37). With N501Y, we observed a reduced capacity for antibodies to neutralize, despite no loss in total-IgG (Figure 2). Similar to N501Y, here we observed using BLI that while L452R did not differ greatly from the ancestral RBD in total IgG binding, it exhibited a stronger affinity (Kd: 27.36 nM; t1.2 : 126.4 s), which in turn could explain the drop in plasma-driven ACE2 inhibition (Supplementary Figure 3; Supplementary Table 4). Likewise, using BLI, we observed that N501T had improved binding affinity (KD: 11.05 nM; t1.2 : 217.8 s) as compared to the ancestral RBD (Supplementary Figure 3; Supplementary Table 4). As such, the combination of poorer IgG binding due loss of epitope and stronger ACE2 affinity for N501T could further impeded ACE2 inhibition by plasma samples. Together, these results suggest that beyond antibody recognition, affinity interactions between RBD and ACE2 exert another layer of influence on the potential for antibodies to effectively block ACE2-binding and achieve neutralization.

### hACE2 polymorphisms alter affinity to SARS-CoV-2 RBD variants

Several ACE2 polymorphisms are present at low frequency within the human population, with certain alleles more prevalent in different populations (Supplementary Table 6). In particular, K26R has been observed at low frequencies across most populations, whereas low frequencies of S19P is predominantly detected within African/African American populations (0.3%) (21, 23). Previous work has shown these polymorphisms confer altered affinity to ancestral RBD, however the binding kinetics to emerged VOC RBD, especially Delta and Omicron, has yet to be described. Here we assessed the binding profiles of three low frequency ACE2 polymorphisms (E35K, K26R and S19P) to 5 VOC RBDs (Alpha, Beta, Delta, Gamma, Omicron (BA.2)) and ancestral RBD using biolayer interferometry (BLI). We found that the polymorphism S19P (KD: 14.38 nM) and K26R (KD: 20.0) demonstrated a 2.04-fold and 1.47 fold increase in binding affinity respectively to ancestral-RBD when compared to WT ACE2 (KD: 28.99 nM) (Figure 4, Supplementary Table 5). Furthermore, this enhanced affinity of the S19P and K26R polymorphisms remains overall elevated against the RBD’s of Beta (KD: S19P=10.51 nM, K26R=17.46 nM), Omicron (BA.2) (KD: S19P=4.97 nM, K26R=5.18 nM) and Delta (KD: S19P= 8.56, K26R=13.47) (Figure 4, Supplementary Table 5). These data suggest that individuals who harbor S19P or K26R ACE2 polymorphisms may have greater susceptibility to infection by SARS-CoV-2 due to increased ACE2 affinity. Contrastingly, ACE2 polymorphism E35K demonstrated weaker binding when compared to ancestral-RBD (KD: 36.63 nM), Omicron (BA.2) (Kd: 18.50 nM) and Delta (Kd: 43.34 nM), suggesting that individuals harboring the E35K ACE2 polymorphism may demonstrate a more protective phenotype due to reduced ACE2 affinity for RBD (Figure 4; Supplementary Table 5). Together, these data suggests that ACE2 polymorphisms, though low in frequency, can modulate binding to ACE2, and by extension may change an individual’s susceptibility to infection.

**Figure 4.**
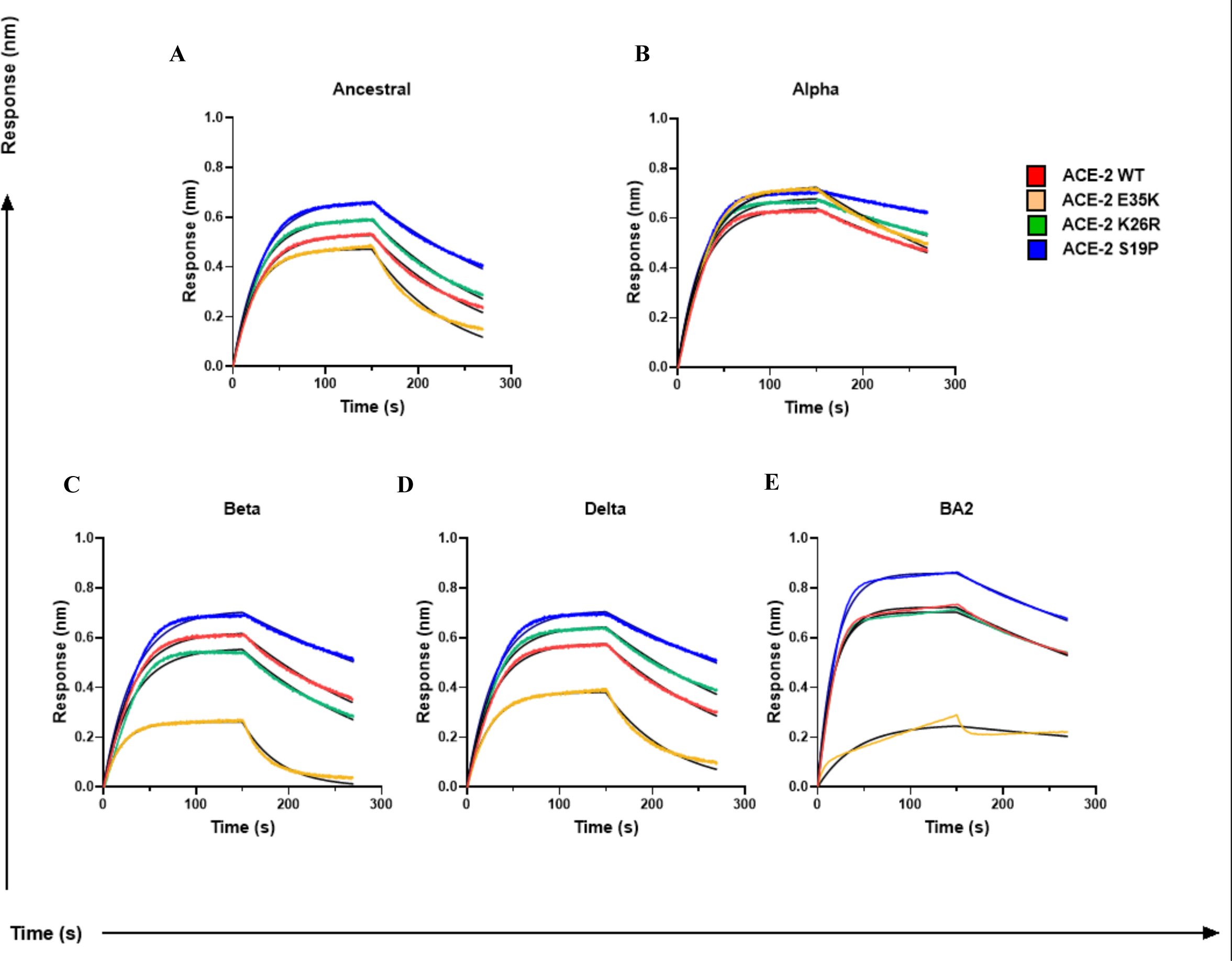
Affinity of hACE2 polymorphisms to VOC RBD’s. Binding profiles of WT hACE2 (purple) and three polymorphisms (E35K, green; K26R, red; S19P, blue) to (A) ancestral-RBD and RBD of variants of concern (B) Alpha (B.1.1.7), (C) Beta (B.1.351), (D) Delta (B.1.617.2), (E) Omicron (BA.2). Sensograms show 100 nM of immobilised RBD of each variant run against 100nm (3ug/ml) ACE2 of each polymorphism to measure binding kinetics.

### Reduced FcγR-binding antibodies to RBD variants

Several previous studies have highlighted an importance for Fc-effector functions during SARS-CoV-2 infection (14,15,17,18,20). Given the observed loss of IgG to certain RBD-variants, we next investigated the impact these mutations conferred on the capacity to for anti-RBD antibodies to engage our FcγRIIa and FcγRIIIa-dimer constructs, a surrogate measure for ADCP and ADCC activity respectively. We used our multiplex panel of 39-RBD to address the impact of these mutations on FcR-binding antibodies as compared to the ancestral RBD. Consistent with our previous observation to total-IgG levels, BNT162b2 vaccinee plasma had a significantly greater capacity (higher reciprocal ED50 vales) to engage both our FcγRIIa and FcγRIIIa-dimer constructs across all variants, in comparison to convalescent plasma (Figure 5A,B). Compared to ancestral-RBD, VOC Beta and Gamma demonstrated a significant decrease in anti-RBD FcγRIIa (9.8 and 14.9-fold-reduction respectively; p < 0.0001) and FcγRIIIa-dimer binding antibodies (8.4, and 7.2-fold-reduction respectively; p < 0.0001) (Figure 5C, D). Likewise, a drop was observed with Omicron BA.2 to both FcγRIIa and FcγRIIIa (3.1 and 3.7-fold reduction respectively; p < 0.0001) (Figure 5E, F). Altogether, our data suggests that the capacity to recruit Fc-effector functions is compromised by mutations found in VOCs (Figure 5; Supplementary Table 3). Furthermore, we observed that single-point mutations with poorer IgG binding, such as N501T and G446S, displayed a significant reduction in FcγR-binding as well (FcγRIIa: 6.2-, 5.0-fold reduction respectively; FcγRIIIa 5.5-, 2.1-fold reduction respectively) (P < 0.0001) (Figure 5C, D; Supplementary Table 3). These results are consistent with the loss of IgG-recognition we observed to our panel of variants (Figure 2). To confirm our findings using the surrogate FcγR dimers, we next measured the capacity for BNT162b2 plasma to induced ADCP using a duplex bead-based assay with a THP- 1 monocyte cell line. FcγRIIa reciprocal ED50 values correlated well to phagocytosis levels of ancestral-RBD by THP-1 monocytes (spearman r = 0.59, p = 0.017), as well as the RBD of Beta (spearman r = 0.52, p = 0.4) (Figure 5G, H). We observed a significant decrease in ADCP activity against ancestral vs Beta RBD (p = 0.0026), supporting the mutations found in VOCs are detrimental to FcγR-binding antibodies (Figure 5I).

**Figure 5.**
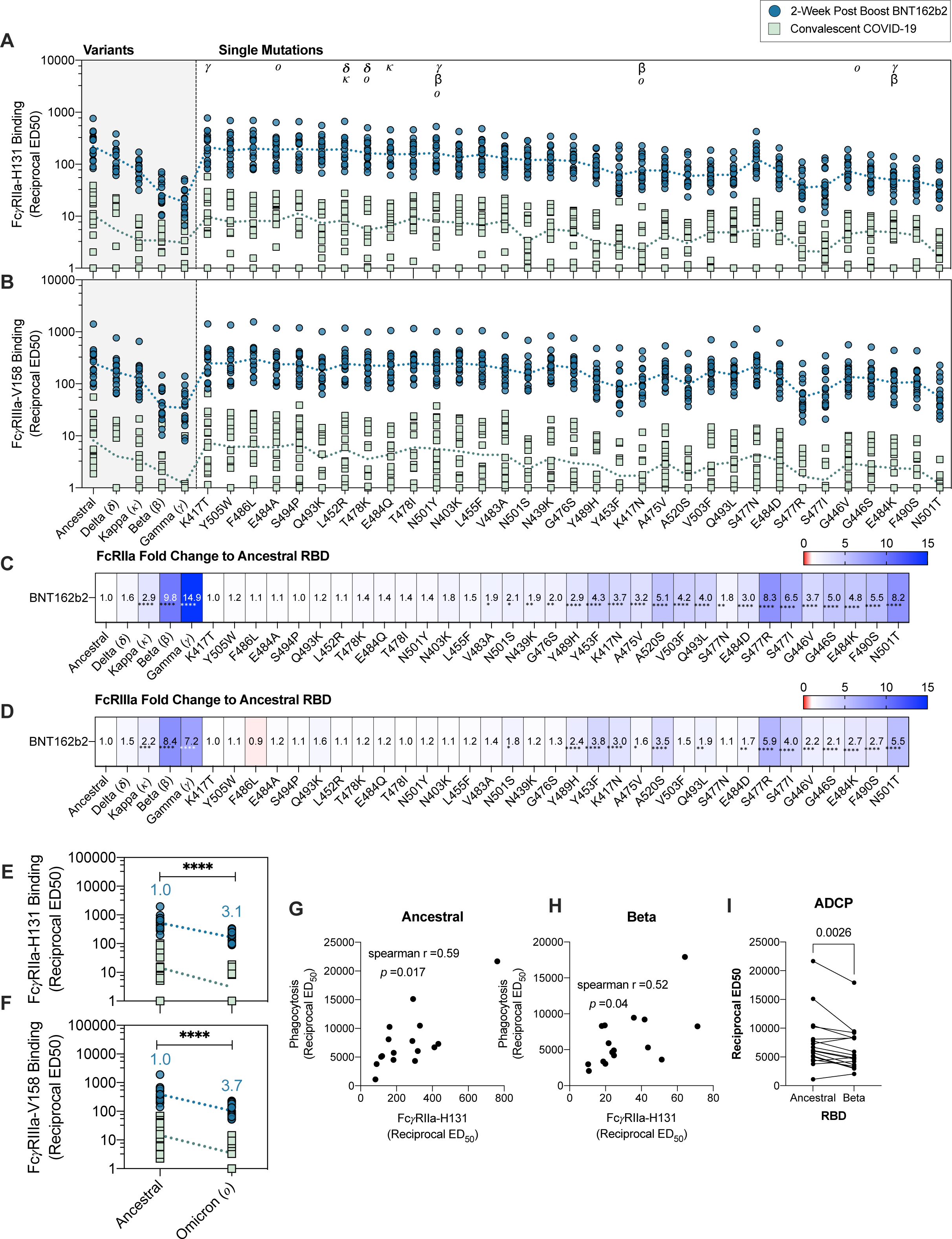
Loss of anti-RBD FcγR-binding antibodies and ADCP responses to VOC RBDs in BNT162b2 and convalescent plasma. (A) FcγRIIa-H131 and (B) FcγRIIIa-V158 binding levels of RBD-specific antibodies using plasma of BNT162b2 (two-weeks following second dose; blue; *n* = 16) and mild/moderate convalescent COVID-19 patients (green; median 38 days post symptom onset; *n* = 15) as measured via multiplexing. (C) Fold change of anti-RBD FcγRIIa-H131 binding levels to RBD-variants in comparison to ancestral-RBD of BNT162b2 plasma. (D) Fold change of FcγRIIIa-V158 levels of RBD variants in comparison to ancestral of BNT162b2 plasma. Reciprocal ED50 values were calculated using the normalized FcγR binding MFI-value for an 8-point serial plasma titration. Fold change was calculated as follows: (geometric mean of reciprocal ED50 or ID50 respectively ancestral-RBD) divided by (geometric mean reciprocal ED50 or ID50 respectively variant-RBD). Friedman’s non-parametric test with Dunn’s multiple comparisons was used to assess statistical significance. (E) *Pan*-IgG binding (Reciprocal ED50) and (F) hACE2 inhibition levels to RBD of Omicron variant with fold-change of BNT162b2 plasma indicated above in blue. Mann-Whitney U-test used to assess statistical significance. (G-H) Spearman correlations of phagocytosis activity (reciprocal ED50) using THP-1 monocytes and multiplex FcRIIa-H131 data (reciprocal ED50) against the ancestral and Beta RBD. (I) ADCP by THP-1 monocytes induced by the plasma of BNT162b2 against the ancestral and Beta RBD shown as phagocytic score ED50s. Wilcoxon matched pairs signed rank test was used to assess statistical significance. *p* < 0.05 (*), *p* < 0.01 (**), *p* < 0.001 (***) and *p* < 0.0001 (****)

## Discussion

The emergence of SARS-CoV-2 variants has led to new waves of infection across the globe, with the RBD of SARS-CoV-2 spike being a key site for escape mutations (1, 40). Here, we showed reduced recognition of RBD-variants by IgG plasma antibodies from both BNT162b2 and convalescent-subjects, with profound loss against VOCs Beta, Gamma, and more recent Omicron, indicative of evolutionary immune escape. Using our surrogate RBD- ACE2 multiplex bead assay, we showed that the drop in IgG-binding against VOCs corresponded to a reduction in neutralization capacity, consistent with others (8,41–43). Furthermore, in line with loss of IgG binding, we also found a decrease in anti-RBD FcγR - binding antibodies against variants, suggesting FcγR functions were not well retained against VOC-RBDs. Interestingly, Bartsch et al., also went on to show that FcγR-functions against the full-VOC spike are less drastically impacted by mutations, likely reflecting the greater number of conserved epitopes harbored on the full-spike that can be targeted by both neutralizing and non-neutralizing antibodies (44).

Consistent with recent literature, all VOC-RBDs (Beta, Gamma, Delta, Omicron (BA.2) bind WT-ACE2 with enhanced affinity, reflecting evolutionary mutations that enhance viral entry and transmission (26, 39). Moreover, a key finding in our study was that human ACE2 polymorphisms, whilst found in low frequencies, also demonstrated distinct variation in affinity to the RBD of VOCs. Here, we provided first insights into the binding affinity of VOC-RBDs, notably Delta and Omicron, to several ACE2 N-terminal polymorphisms – the site for RBD docking. K26R (rs4646116), – the second most frequent human ACE2 polymorphism (0.4% allele frequency), displayed enhanced binding affinity to ancestral-RBD, and VOCs, Alpha, Delta, while maintaining similar affinity to Omicron (24, 45). S19P which is predominantly observed within African/African America populations (0.3%) also showed enhanced affinity to ancestral RBD, along with all tested VOC (Alpha, Beta, Delta, and Omicron) (24, 45). These stronger binding affinities to VOCs RBDs may pre-dispose subsets of individuals harboring these polymorphisms to become more susceptible to breakthrough infections by SARS-CoV-2 variants (21,22,24,45–47). Intriguingly, K26R (rs4646116)– demonstrated slightly weaker binding to RBD-Beta. E35K (rs1348114695, which has been predominantly detected at very low frequencies (0.01%) in East Asian populations) also had decreased affinity to ancestral RBD and more importantly demonstrated an even greater loss in binding affinity to VOC RBDs, particularly Beta, Delta, and Omicron, which may confer some level of natural protection against breakthrough infections by these variants. Our work suggests that large genetic linkage studies together with VOC infection and severity data could be informative.

Assessing single point mutations found in the RBD can provide valuable insights into the importance of these mutations on overall viral fitness. Here, we found numerous point mutations imparted little impact on anti-RBD IgG-recognition as well as FcγR-binding engagement. This was observed for RBD point mutations L452R and T478K (both found in Delta), which may account for the limited reduction in total IgG binding and FcγR-binding observed against Delta RBD. However, we noted that L452R-RBD had a longer binding half-life to WT-ACE2, which could contribute to the observed loss of ACE2 neutralization. Indeed, it has been described that L452R increases SARS-CoV-2 infectivity and fusogenicity, thus may confer an evolutionary advantage previously observed with Delta and more recently the Omicron sublineages BA.4 and BA.5 (48). On the other hand, certain point mutations, such as G446S (found in Omicron), G446V, S477I and S477R conferred reduced recognition by IgG. Therefore, suggesting mutations at amino acid position 446 and 477 are advantageous for viral immune escape, which is further reinforced by loss of FcγR-binding. Interestingly, while S477N (found in Omicron) also resulted in a drop in total IgG binding, it was less disruptive to FcγR-dimer engagement than both S477I and S477R (24, 25). We also observed the largest fall in total IgG binding against N501T, which displayed increased ACE2 affinity and longer binding half-life resulting in a significant loss in neutralization. While N501T had weaker ACE2-binding affinity compared to N501Y – a key RBD mutation found in Delta, Beta and now Omicron – the loss in FcγR-binding was more profound with N501T, corresponding to the drastic reduction in total IgG binding. This is of growing concern for zoonotic transmission as N501T is a dominant RBD mutation identified in SARS-CoV-2-infected minks, ferrets, and deer (49–51).

Several future studies are suggested by our work. While our cohort of BNT162b2- individuals received two-doses of vaccine, the current approved regimen is now expanded to three-doses in Australia and many other countries. Three doses enhance antibody responses to VOCs overall and it will be important to dissect particular mutations for their impact on improved RBD-specific antibody responses. It is also unclear how our finding observed with BNT162b2 -vaccination will translate to other SARS-CoV-2 vaccines with more or less potent responses (such as mRNA-1273, adenoviral vectors, inactivated viruses, and protein subunit vaccines). Furthermore, neutralizing antibody responses elicited from current Omicron BA.2 infections have been shown to differ from that of earlier waves (Omicron BA.1 and Delta) and this will also likely impact the specificity of RBD responses (52). Lastly, due to resource constraints, we opted to use commercially available Omicron BA.2 and WT RBD to study responses against Omicron. The subtle variations between our in-house and the commercially available RBD expression templates led to noticeable differences in multiplex assay sensitivity.

Overall, our data show that RBD mutations, both common point mutations as well as combinations of mutations in VOCs, not only have an impact on affinity to receptor ACE2, but also IgG-binding and the Fc functional antibody response, thereby providing an important site for viral adaptions.

## Data Availability

All data produced in the present work are contained in the manuscript

## Acknowledgements

This study was supported by the Victorian Government and Medical Research Future Fund (MRFF) GNT2002073 (to A.K.W, P.M.H., W.-H.T., D.I.G., S.J.K., and A.W.C.), GNT2005544 (to A.K.W., J.A.J., D.I.G., S.J.K., and A.W.C), the Paul Ramsay Foundation (A.K.W., D.I.G., S.J.K. and A.W.C.). A.K.W, J.A.J., D.I.G., W.-H.T., S.J.K. and A.W.C. are supported by NHMRC fellowships. W-H.T. is a Howard Hughes Medical Institute–Wellcome Trust International Research Scholar (208693/Z/17/Z). N.A.G. is supported by an ARC DECRA fellowship.

## Authors contributions

E.R.H., S.K.D., K.J.S., and A.W.C drafted the original manuscript. E.R.H, S.K.D., P.R., E.L., K.J.S. and A.W.C. designed experimental protocols. E.R.H, S.K.D., P.R., E.L., R.A.P., K.J.S. performed experiments and analyzed data. L.T., P.P, B.D.W., P.M.H., A.K.W, S.R., N.A.G., D.I.G. and W-H.T contributed unique reagents. J.A.J. and S.J.K. contributed unique samples. K.J.S., S.J.K. and A.W.C. conceived and supervised the study. All authors reviewed the manuscript.

## Data and materials availability

All relevant data is available in the manuscript or in the supplementary materials. Any additional supporting data can be provided upon request.

## Supplementary Figure Legends

**Supplementary Figure 1. Schematic representation of duplex phagocytosis assay and gating strategy.** (A) Schematic of the antibody dependent phagocytosis (ADCP) duplex assay. Briefly, APC fluorescent beads coated with ancestral RBD, while FITC fluorescent beads coated with RBD Beta were added to wells and incubated with plasma for 2 hours at 37°C. Following incubation, THP-1 monocytes were added to wells and incubated with opsonised beads for 2 hours under cell culture conditions before fixing cells for flow cytometry. (B) Representative gating strategy using pooled 2-week post vaccination plasma (blue), pooled baseline plasma (red) and no plasma (grey). Gating was performed by gating on THP-1 monocytes, single cells and finally cells positive for RBDWT beads and RBD Beta beads. (C) Visualisation of single and double bead positive cells as dot plots for representative samples.

**Supplementary Figure 2. IgG subclass recognition of SARS-CoV-2 antigens and correlations to FcγR engagement.** Plasma antibody isotype response (pan IgG, IgA1, IgM) (A-C), IgG subclass responses (IgG1, IgG2, IgG3 IgG4) (D-G) and FcγR engagement (FcγR3av and FcγR2aH) (H-I) to NP, RBD, S1 and spike trimer were profiled via multiplex. Convalescent mild/moderate COVID-19 patients (green square; median 38 days post symptom onset; *n* = 15) and BNT162b2-vaccinated individuals at baseline (purple triangle; *n* = 16) and two-weeks following second dose (blue circle; *n* = 16) were assessed. (J) Pearson R correlation matrix of IgG subclass binding (IgG1, IgG2, IgG3, IgG4) and FcγR engagement (FcγR3av and FcγR2aH) with R values in bold and *P*-values described in italics.

**Supplementary Figure 3. Affinity of wild-type (WT) hACE2 to the RBD mutations.** Sensograms of WT-human ACE2 (hACE2) binding affinity to (A) K417N, (B) K417T, (C) G446S, (D) L452R (E) L452R E484Q, (F) Y453F, (G) S477N, (H) T478K, (I) E484D, (J) E484K and (K) 484Q, (L) S494P and (M) N501T as measured by biolayer interferometry (BLI). Binding of 2-fold serial dilutions of each SARS-CoV-2 RBD variant (100 nM (red), 50 nM (yellow), 25 nM (green), 12 nM (blue), and 6 nM (pink)) was measured to immobilised hACE2 (3ug/ml). Binding curves representative of 2 independent experiments for each variant are plotted (solid-colored lines), globally fitted to a 1:1 binding model (black line).

**Supplementary Table 1. Antigens used in SARS-CoV-2-specific array**

**Supplementary Table 2. RBD recombinant proteins (variants and single point mutations) used in RBD array**

**Supplementary Table 3. Friedmans Test using Dunn’s multiple comparisons statistical values for comparison of IgG-binding, ACE2-inhibition, FcRIIa-binding and FcRIIIa- binding of RBD-variants to ancestral RBD**

**Supplementary Table 4. Affinity constants of wild-type (WT) hACE2 to RBD with single point mutations.** Measured via BLI, calculated from 2 independent experiments

**Supplementary Table 5. Affinity of human ACE2 polymorphisms to SARS-CoV-2 RBD Variants**. Measured via BLI, calculated from 2 independent experiments

**Supplementary Figure 6. Frequency of human ACE2 polymorphisms (E35K, K26R, S19P) found within human populations.** Polymorphism data collated from Genome Aggregation Database (gnomAD; https://gnomad.broadinstitute.org)

