## Supplementary material for "Altered affinity to ACE2 and reduced Fc functional antibodies to SARS-CoV-2 RBD variants": Supp materials

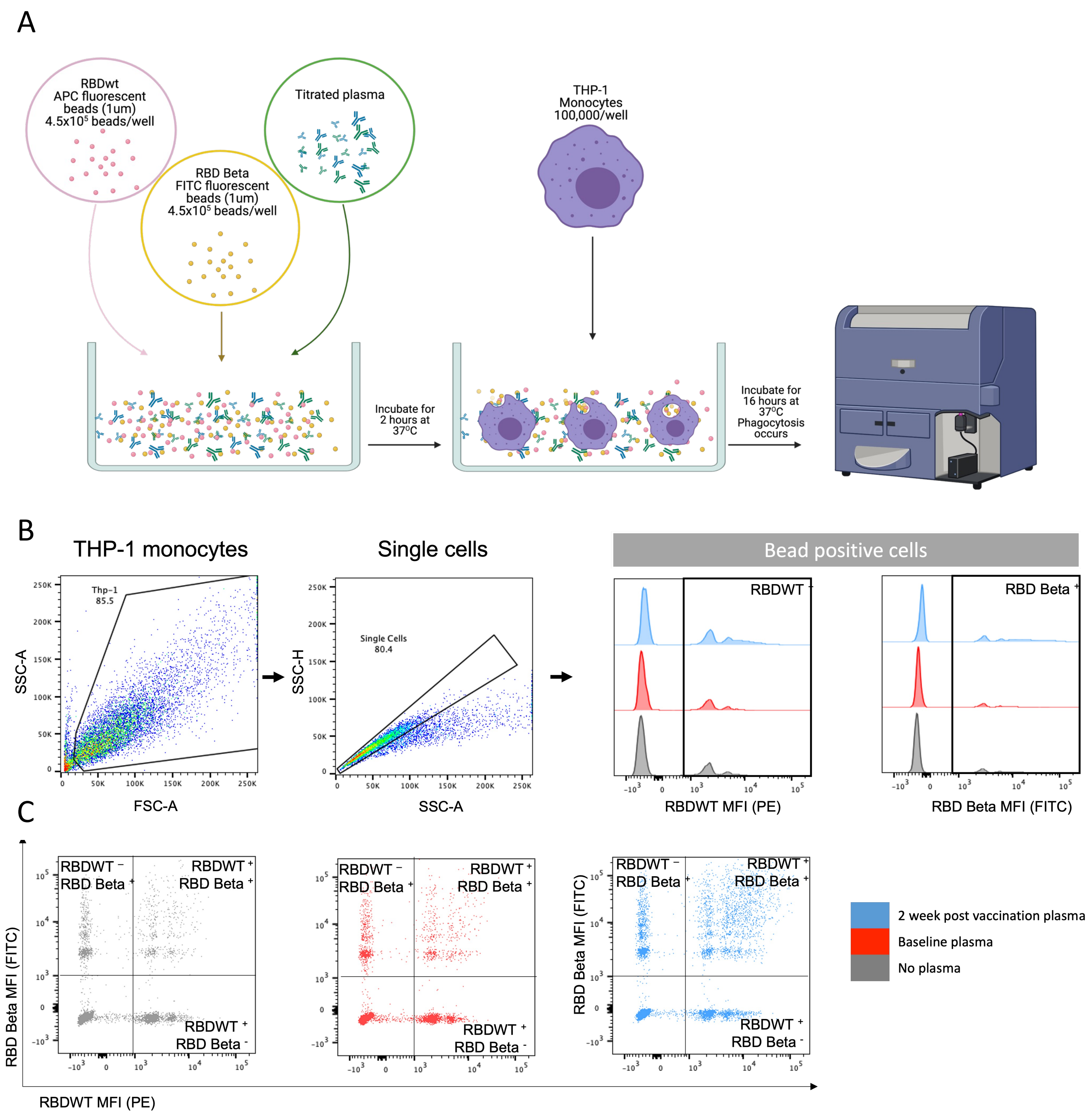


**Supplementary Figure 1**

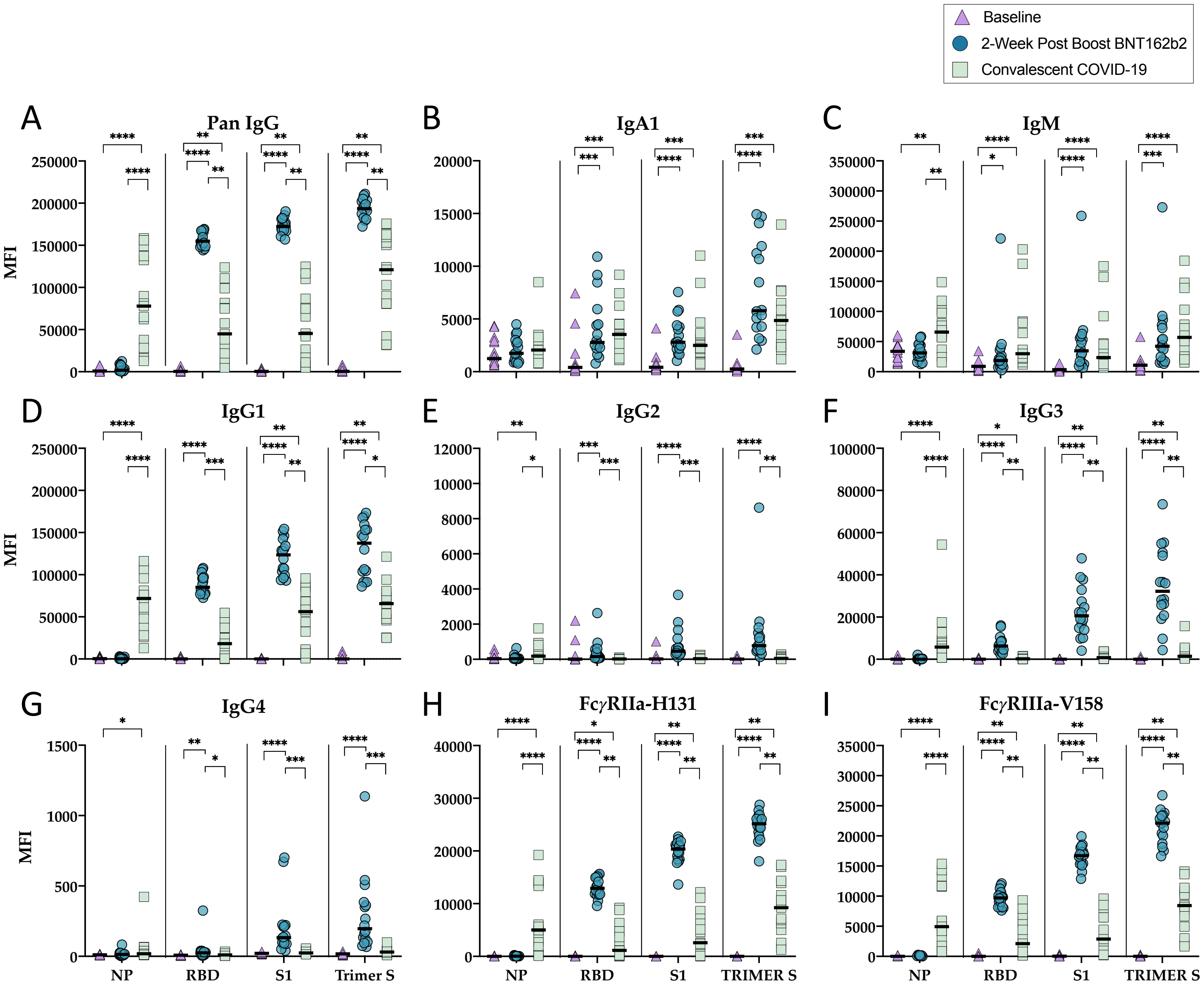


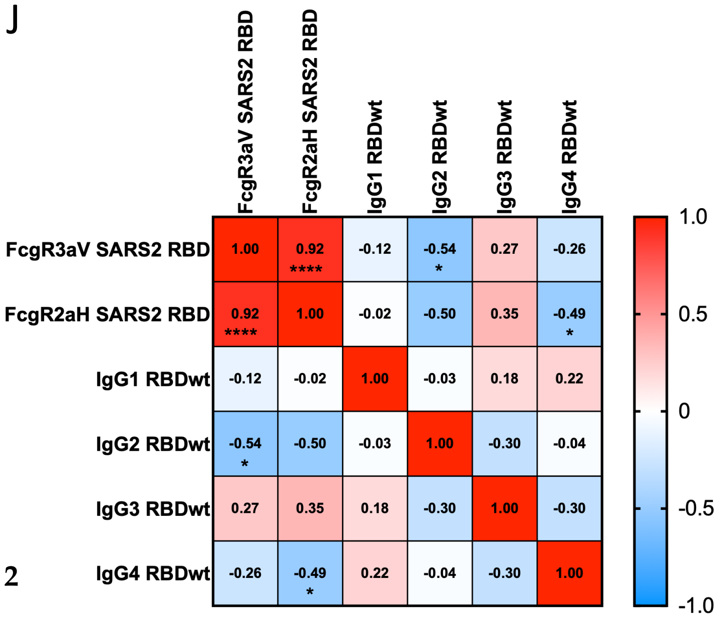


Supplementary Figure 2

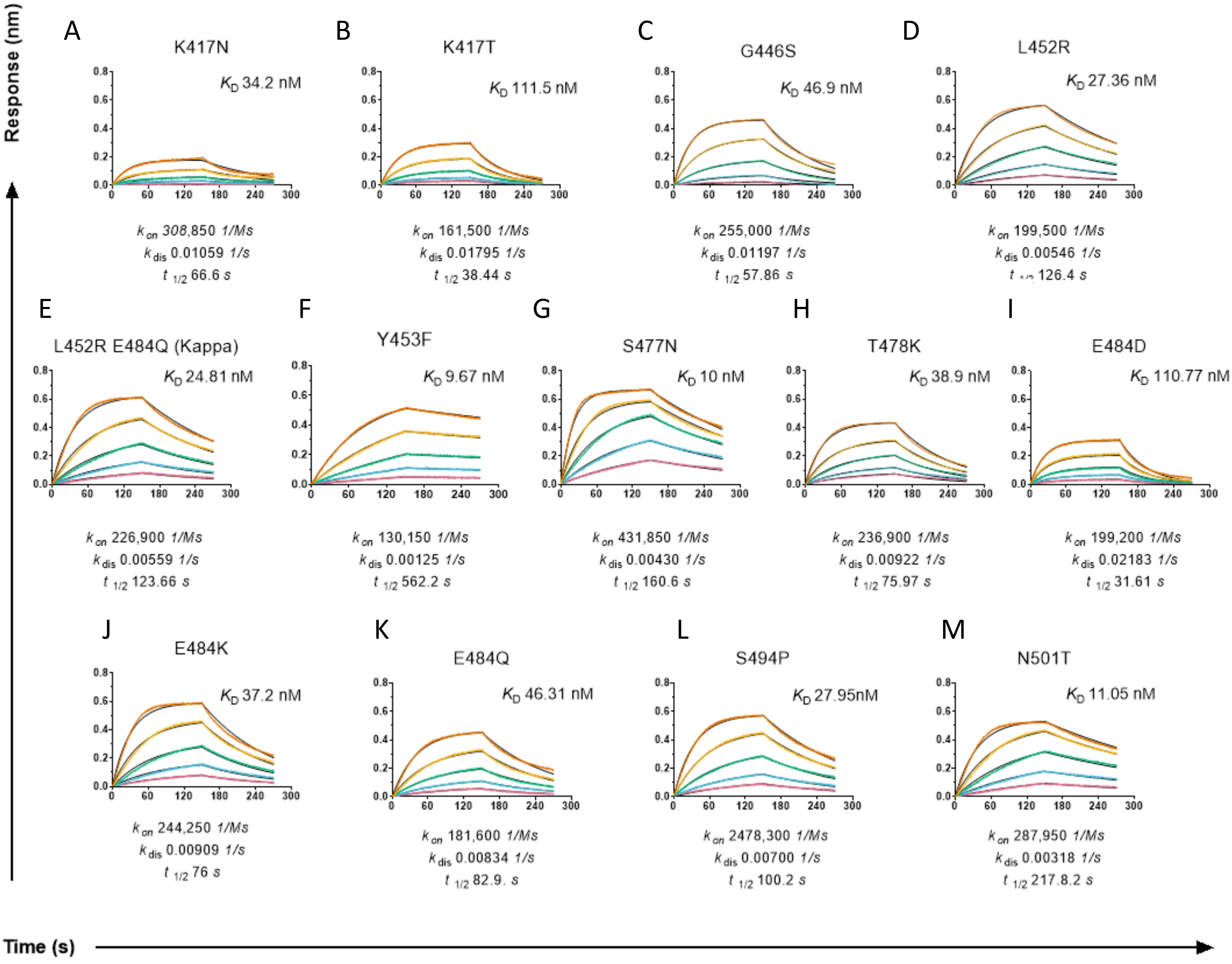


**Supplementary Figure 3**

**Supplementary Table 1**

| Bead Supplier | Bead Region | Bead Catalogue Number | Antigen | Protein Coupled/1x10^7 beads |
|---|---|---|---|---|
| Biorad | 14 | MC10014-01 | Spike S1 | 100ug |
| Biorad | 22 | MC10022-01 | Nucleoprotein | 40ug |
| Biorad | 76 | MC10076-01 | Spike Trimer | 100ug |
| Biorad | 74 | MC10074-01 | Influenza H1Cal2009 (control) | 100ug |
| Biorad | 15 | MC10015-01 | SIVgp120 (control) | 100ug |

### Supplementary Table 2

| Bead Supplier | Bead Region | Bead Catalogue Number | RBD Recombinant Protein | Protein Coupled/1x10^7 beads |
|---|---|---|---|---|
| Biorad | 6 | MC12006-01 | E406W | 100ug |
| Biorad | 7 | MC10007-01 | K417N | 100ug |
| Biorad | 9 | MC10009-01 | Ancestral wild-type (WT) | 100ug |
| Biorad | 12 | MC10012-01 | Y453F | 100ug |
| Biorad | 13 | MC10013-01 | R403K | 100ug |
| Biorad | 15 | MC10015-01 | G446S | 100ug |
| Biorad | 16 | MC12016-01 | G446V | 100ug |
| Biorad | 18 | MC10018-01 | L455F | 100ug |
| Biorad | 20 | MC10020-01 | A475V | 100ug |
| Biorad | 22 | MC10022-01 | G476S | 100ug |
| Biorad | 25 | MC10025-01 | S477I | 100ug |
| Biorad | 26 | MC10026-01 | S477N | 100ug |
| Biorad | 29 | MC10029-01 | N439K | 100ug |
| Biorad | 30 | MC10030-01 | T478I | 100ug |
| Biorad | 33 | MC10033-01 | N501T | 100ug |
| Biorad | 34 | MC10034-01 | G485R | 100ug |
| Biorad | 35 | MC10035-01 | Q493K | 100ug |
| Biorad | 37 | MC10037-01 | S494L | 100ug |
| Biorad | 36 | MC10036-01 | V483A | 100ug |
| Biorad | 38 | MC10038-01 | E484A | 100ug |
| Biorad | 39 | MC10039-01 | E484D | 100ug |
| Biorad | 42 | MC10042-01 | E484K | 100ug |
| Biorad | 44 | MC10044-01 | F490S | 100ug |
| Biorad | 46 | MC10045-01 | N501S | 100ug |
| Biorad | 47 | MC10047-01 | S477R | 100ug |
| Biorad | 48 | MC10048-01 | Y505W | 100ug |
| Biorad | 50 | MC12050-01 | F486L | 100ug |
| Biorad | 51 | MC10051-01 | Q493L | 100ug |
| Biorad | 52 | MC10052-01 | S494P | 100ug |
| Biorad | 53 | MC10053-01 | N501Y | 100ug |
| Biorad | 55 | MC10055-01 | T478K | 100ug |
| Biorad | 61 | MC10061-01 | N501Y, E484K, K417N (B.1.251; Beta) | 100ug |
| Biorad | 62 | MC10062-01 | V503F | 100ug |
| Biorad | 64 | MC10064-01 | K417T | 100ug |
| Biorad | 70 | MC10070-01 | L452R | 100ug |
| Biorad | 72 | MC10072-01 | E484Q | 100ug |
| Biorad | 73 | MC10073-01 | N501Y, E484K, K417T (P1; Gamma) | 100ug |
| Biorad | 74 | MC10074-01 | S477N | 100ug |
| Biorad | 75 | MC10075-01 | Y489H | 100ug |
| Biorad | 76 | MC10076-01 | L452R E484Q (B.1.617.1; Kappa) | 100ug |
| Biorad | 77 | MC10077-01 | A520S | 100ug |
| Biorad | 78 | MC10078-01 | L452R, T478K (B.1.617.2; Delta) | 100ug |
| Biorad | 22 | MC10078-22 | Ancestral RBD (SinoBiological; Catalogue # 40592-V08H) | 100ug |
| Biorad | 55 | MC10078-55 | Omicron RBD BA.2 (SinoBiological; Catalogue #40592-V08H123) | 100ug |

### Supplementary Table 3

| Dunn's multiple comparisons test | IgG Rank sum diff. | IgG Summary | IgG Adjusted P Value | ACE Rank sum diff. | ACE Summary | ACE Adjusted P Value | FcR2a Rank sum diff. | FcR2a Summary | FcR2a Adjusted P Value | FcR3a Rank sum diff. | FcR3a Summary | FcR3a Adjusted P Value |
|---|---|---|---|---|---|---|---|---|---|---|---|---|
| **Ancestral vs. Delta** | 156 | ns | 0.4841 | 161.5 | ns | 0.3774 | 197 | ns | 0.0639 | 199 | ns | 0.0573 |
| **Ancestral vs. Kappa** | 315 | **** | <0.0001 | 139.5 | ns | 0.9799 | 359 | **** | <0.0001 | 290 | *** | 0.0001 |
| **Ancestral vs. Beta** | 510 | **** | <0.0001 | 327.5 | **** | <0.0001 | 536 | **** | <0.0001 | 465 | **** | <0.0001 |
| **Ancestral vs. Gamma** | 546 | **** | <0.0001 | 352.5 | **** | <0.0001 | 550 | **** | <0.0001 | 464 | **** | <0.0001 |
| **Ancestral vs. K417T** | -38 | ns | >0.9999 | -172.5 | ns | 0.2246 | 3 | ns | >0.9999 | 18.5 | ns | >0.9999 |
| **Ancestral vs. Y505W** | -23 | ns | >0.9999 | 272.5 | *** | 0.0005 | 80 | ns | >0.9999 | 0 | ns | >0.9999 |
| **Ancestral vs. F486L** | 19 | ns | >0.9999 | -204.5 | * | 0.0423 | 33 | ns | >0.9999 | -92 | ns | >0.9999 |
| **Ancestral vs. E484A** | 21 | ns | >0.9999 | 101 | ns | >0.9999 | 64 | ns | >0.9999 | 31 | ns | >0.9999 |
| **Ancestral vs. S494P** | 65.5 | ns | >0.9999 | 139.5 | ns | 0.9799 | 52 | ns | >0.9999 | 31 | ns | >0.9999 |
| **Ancestral vs. Q493K** | 43 | ns | >0.9999 | -146.5 | ns | 0.7321 | 52 | ns | >0.9999 | 182.5 | ns | 0.1367 |
| **Ancestral vs. L452R** | 47.5 | ns | >0.9999 | 204.5 | * | 0.0423 | 34 | ns | >0.9999 | -6 | ns | >0.9999 |
| **Ancestral vs. T478K** | 78 | ns | >0.9999 | -121.5 | ns | >0.9999 | 105 | ns | >0.9999 | 52 | ns | >0.9999 |
| **Ancestral vs. E484Q** | 120 | ns | >0.9999 | -5.5 | ns | >0.9999 | 135 | ns | >0.9999 | 59 | ns | >0.9999 |
| **Ancestral vs. T478I** | 140 | ns | 0.9601 | -51.5 | ns | >0.9999 | 137 | ns | >0.9999 | -12 | ns | >0.9999 |
| **Ancestral vs. N501Y** | 107 | ns | >0.9999 | 359.5 | **** | <0.0001 | 118 | ns | >0.9999 | 28 | ns | >0.9999 |
| **Ancestral vs. N403K** | 152 | ns | 0.5776 | 54.5 | ns | >0.9999 | 199 | ns | 0.0573 | -16 | ns | >0.9999 |
| **Ancestral vs. L455F** | 173 | ns | 0.2192 | -39 | ns | >0.9999 | 145 | ns | 0.78 | 30 | ns | >0.9999 |
| **Ancestral vs. V483A** | 221 | * | 0.0162 | -29.5 | ns | >0.9999 | 210.5 | * | 0.0301 | 131 | ns | >0.9999 |
| **Ancestral vs. N501S** | 224 | * | 0.0136 | -179 | ns | 0.1631 | 224.5 | * | 0.0132 | 223 | * | 0.0144 |
| **Ancestral vs. N439K** | 232.5 | ** | 0.008 | 204 | * | 0.0434 | 236 | ** | 0.0064 | 28 | ns | >0.9999 |
| **Ancestral vs. G476S** | 246 | ** | 0.0034 | 14 | ns | >0.9999 | 258 | ** | 0.0015 | 106.5 | ns | >0.9999 |
| **Ancestral vs. Y489H** | 276 | *** | 0.0004 | -36.5 | ns | >0.9999 | 305 | **** | <0.0001 | 324 | **** | <0.0001 |
| **Ancestral vs. Y453F** | 284 | *** | 0.0002 | 282.5 | *** | 0.0003 | 397 | **** | <0.0001 | 385 | **** | <0.0001 |
| **Ancestral vs. K417N** | 256 | ** | 0.0017 | -107.5 | ns | >0.9999 | 357 | **** | <0.0001 | 336 | **** | <0.0001 |
| **Ancestral vs. A475V** | 311 | **** | <0.0001 | 14.5 | ns | >0.9999 | 350 | **** | <0.0001 | 211 | * | 0.0292 |
| **Ancestral vs. A520S** | 354 | **** | <0.0001 | -133.5 | ns | >0.9999 | 421 | **** | <0.0001 | 381 | **** | <0.0001 |
| **Ancestral vs. V503F** | 367 | **** | <0.0001 | 6.5 | ns | >0.9999 | 404 | **** | <0.0001 | 177 | ns | 0.1802 |
| **Ancestral vs. Q493L** | 360 | **** | <0.0001 | 301.5 | **** | <0.0001 | 404 | **** | <0.0001 | 247 | ** | 0.0032 |
| **Ancestral vs. S477N** | 394 | **** | <0.0001 | 147 | ns | 0.7167 | 230 | ** | 0.0094 | 51 | ns | >0.9999 |
| **Ancestral vs. E484D** | 428 | **** | <0.0001 | 212.5 | * | 0.0268 | 339 | **** | <0.0001 | 242.5 | ** | 0.0042 |
| **Ancestral vs. S477R** | 408.5 | **** | <0.0001 | 7.5 | ns | >0.9999 | 516 | **** | <0.0001 | 450 | **** | <0.0001 |
| **Ancestral vs. S477I** | 428.5 | **** | <0.0001 | -91 | ns | >0.9999 | 492 | **** | <0.0001 | 415 | **** | <0.0001 |
| **Ancestral vs. G446V** | 439.5 | **** | <0.0001 | 131.5 | ns | >0.9999 | 344 | **** | <0.0001 | 282 | *** | 0.0003 |
| **Ancestral vs. G446S** | 465 | **** | <0.0001 | 187.5 | ns | 0.1058 | 440 | **** | <0.0001 | 305 | **** | <0.0001 |
| **Ancestral vs. E484K** | 490 | **** | <0.0001 | 119.5 | ns | >0.9999 | 467 | **** | <0.0001 | 365 | **** | <0.0001 |
| **Ancestral vs. F490S** | 502 | **** | <0.0001 | -42.5 | ns | >0.9999 | 483 | **** | <0.0001 | 351 | **** | <0.0001 |
| **Ancestral vs. N501T** | 495 | **** | <0.0001 | 260.5 | ** | 0.0013 | 507 | **** | <0.0001 | 447 | **** | <0.0001 |

**Supplementary Table 4.**

| RBD | $K_D$ (nM) | SD | $k_{on}$ ($\mu M^{-1}s^{-1}$) | SD | $k_{off}$ ($s^{-1}$) | SD |
|---|---|---|---|---|---|---|
| **Ancestral** | 29.4 | 2.62 | 0.28 | 0.019 | 0.00825 | 0.00016 |
| **Alpha (N501Y)** | 6.4 | 0.49 | 0.33 | 0.003 | 0.0021 | 0.00018 |
| **Beta** | 14.6 | 2.28 | 0.29 | 0.013 | 0.00427 | 0.00086 |
| **Delta** | 21.5 | 0.63 | 0.22 | 0.006 | 0.00480 | 0.00001 |
| **Gamma** | 15.0 | 1.64 | 0.22 | 0.011 | 0.00325 | 0.00052 |
| **Omicron BA2** | 11.0 | 0.03 | 0.39 | 0.02 | 0.00427 | 0.00019 |
| **K417N** | 34.20 | 5.80 | 0.31 | 0.013 | 0.01059 | 0.00226 |
| **K417T** | 111.50 | 12.02 | 0.16 | 0.011 | 0.01795 | 0.00078 |
| **G446S** | 46.9 | 3.18 | 0.26 | 0.003 | 0.01197 | 0.00097 |
| **L452R** | 27.36 | 0.08 | 0.20 | 0.004 | 0.00546 | 0.00010 |
| **L452R E484Q (Kappa)** | 24.81 | 3.77 | 0.23 | 0.019 | 0.00559 | 0.00039 |
| **Y453F** | 9.67 | 0.42 | 0.13 | 0.031 | 0.00125 | 0.00024 |
| **S477N** | 10.0 | 0.18 | 0.43 | 0.034 | 0.0043 | 0.00026 |
| **T478K** | 38.9 | 6.33 | 0.24 | 0.002 | 0.00922 | 0.00160 |
| **E484D** | 110.77 | 16.88 | 0.2 | 0.0281 | 0.02183 | 0.00024 |
| **E484K** | 37.2 | 1.56 | 0.24 | 0.001 | 0.00909 | 0.00033 |
| **E484Q** | 46.31 | 7.62 | 0.18 | 0.020 | 0.00834 | 0.0005 |
| **S494P** | 27.95 | 2.62 | 0.25 | 0.029 | 0.00689 | 0.00016 |
| **N501T** | 11.1 | 1.10 | 0.29 | 0.01 | 0.00318 | 0.00023 |

$K_D$ = dissociation constant, $k_{on}$ = association rate constant, $k_{off}$ = dissociation rate constant
Measured via BLI, calculated from 2 independent experiments

**Supplementary Table 5.**

| RBD | hACE-2 variant | Conc. (nM) | $K_D$ (nM) | SD | Ratio to WT ACE2 | $k_{on}$ ($\mu M^{-1}s^{-1}$) | SD | $k_{off}$ ($s^{-1}$) | SD |
|---|---|---|---|---|---|---|---|---|---|
| Ancestral | WT | 100 | 29.40 | 2.6 | | 0.2800 | 0.019 | 0.0083 | 0.00016 |
| | E35K | 100 | 36.63 | 3.6 | 0.80 | 0.3013 | 0.007 | 0.0110 | 0.00085 |
| | K26R | 100 | 20.00 | 3.4 | 1.47 | 0.3074 | 0.025 | 0.0061 | 0.00054 |
| | S19P | 100 | 14.38 | 1.2 | 2.04 | 0.2844 | 0.002 | 0.0041 | 0.00038 |
| Alpha (N501Y) | WT | 100 | 6.4 | 0.49 | | 0.3300 | 0.003 | 0.0021 | 0.00018 |
| | E35K | 100 | 9.23 | 3.71 | 0.69 | 0.3002 | 0.014 | 0.0027 | 0.00098 |
| | K26R | 100 | 4.97 | 1.81 | 1.29 | 0.3404 | 0.013 | 0.0017 | 0.00055 |
| | S19P | 100 | 2.93 | 0.90 | 2.19 | 0.3322 | 0.008 | 0.0010 | 0.00032 |
| Beta | WT | 100 | 14.60 | 2.3 | | 0.2900 | 0.013 | 0.0043 | 0.00086 |
| | E35K | 100 | 72.52 | 29.6 | 0.20 | 0.2832 | 0.017 | 0.0203 | 0.00713 |
| | K26R | 100 | 17.49 | 7.9 | 0.84 | 0.3256 | 0.093 | 0.0053 | 0.00093 |
| | S19P | 100 | 10.51 | 0.9 | 1.39 | 0.2627 | 0.026 | 0.0028 | 0.00005 |
| Delta | WT | 100 | 21.50 | 0.6 | | 0.2200 | 0.006 | 0.0048 | 0.00001 |
| | E35K | 100 | 43.34 | 12.1 | 0.50 | 0.3062 | 0.049 | 0.0130 | 0.00156 |
| | K26R | 100 | 13.47 | 2.5 | 1.60 | 0.3295 | 0.042 | 0.0044 | 0.00025 |
| | S19P | 100 | 8.56 | 1.4 | 2.51 | 0.3215 | 0.026 | 0.0027 | 0.00022 |
| Omicron BA.2 | WT | 100 | 11.00 | 0.0 | | 0.3900 | 0.020 | 0.0043 | 0.00019 |
| | E35K | 100 | 18.50 | 5.8 | 0.59 | 0.2817 | 0.018 | 0.0052 | 0.00131 |
| | K26R | 100 | 5.18 | 1.6 | 2.12 | 0.6210 | 0.075 | 0.0033 | 0.00139 |
| | S19P | 100 | 4.97 | 1.2 | 2.21 | 0.5631 | 0.077 | 0.0028 | 0.00106 |

Measured via BLI, calculated from 2 independent experiments

**Supplementary Table 6**

| Population | Ashkenazi Jewish | European (non-Finnish) | Other | Latino/Admixed American | South Asian | African/African American | European (Finnish) | East Asian |
|---|---|---|---|---|---|---|---|---|
| ***K26R Allele Frequency*** | 0.01188 | 0.005868 | 0.003378 | 0.003249 | 0.001313 | 0.000946 | 0.000483 | 0.000067 |
| ***S19P Allele Frequency*** | 0 | 0 | 0.0001882 | 0 | 0 | 0.003323 | 0 | 0 |
| ***E35K Allele Frequency*** | 0 | 0.000012 | 0 | 0 | 0 | 0 | 0 | 0.000144 |

Polymorphism data collated from Genome Aggregation Database (gnomAD v2.1.1; https://gnomad.broadinstitute.org)
